# Five Days of Acute Intermittent Hypoxia Is Safe and Improves Somatic Motor Function in Persons with Multiple Sclerosis

**DOI:** 10.64898/2026.11.09.26367650

**Authors:** Aravind Nehrujee, Rachel Kalvakota, Alexander Barry, Leah O’Shea, Hannah R. Johnson, Chendong Zou, Jordan N. Northrop, Kailynn Mannella, Amy Zhou, Weena Dee, William Z. Rymer, Alexander J. Nemeth, Bruce A. Cohen, Molly G. Bright, Milap S. Sandhu

**Affiliations:** Shirley Ryan AbilityLab, Chicago, IL, United States; Department of Physical Medicine and Rehabilitation, Northwestern University Feinberg School of Medicine, Chicago, IL, United States; Department of Biomedical Engineering, McCormick School of Engineering and Applied Sciences, Northwestern University, Evanston, IL, United States; Department of Physical Therapy and Human Movement Sciences, Feinberg School of Medicine, Northwestern University, Chicago, IL, United States; Ken & Ruth Davee Department of Neurology, Northwestern University Feinberg School of Medicine, Chicago, IL, United States; Department of Radiology, Northwestern University Feinberg School of Medicine, Chicago, IL, United States

**Author notes:** **Corresponding author:** Milap S. Sandhu, PT, PhD, Shirley Ryan AbilityLab, 355 East Erie Street, Chicago, IL 60611, United States. **Trial registration:** ClinicalTrials.gov NCT06276634.

**Keywords:** acute intermittent hypoxia, multiple sclerosis, neuroplasticity, muscle strength, walking, rehabilitation

## Abstract

Walking impairment affects up to 75% of persons with multiple sclerosis (MS), and the benefits of rehabilitation are constrained by limited capacity for training-induced neuroplasticity. Acute intermittent hypoxia (AIH), brief repeated exposures to low inspired oxygen, triggers serotonin-dependent spinal plasticity that enhances motor output. In spinal cord injury, daily AIH paired with walking training has produced larger walking gains than training alone, positioning AIH as a promising primer for rehabilitation. Before AIH can be combined with training in MS, its safety and stand-alone effects with repeated daily dosing must be established in a population with demyelinating disease. We therefore conducted the first multi-day AIH trial in persons with MS to characterize safety and tolerability and to evaluate preliminary effects on ankle strength and walking.

**METHODS:** Twenty-three ambulatory adults with MS (Expanded Disability Status Scale [EDSS] 3.0-6.5; 15 relapsing-remitting, 8 secondary-progressive) completed a randomized, double-masked, placebo-controlled crossover trial. Participants received AIH (15 × ∼60 s episodes; FiO_2_ = 0.09; SpO_2_ target 84-88%) and Sham in a counterbalanced order, separated by a ≥4-week washout. Assessments at pre-intervention, post-intervention, and follow-up included ankle plantarflexion and dorsiflexion torque, 6-Minute Walk Test (6MWT), Timed 25-Foot Walk (T25FW), and MS Walking Scale-12 (MSWS-12). Treatment effects were estimated using analysis of covariance (ANCOVA) with participant as a random effect.

**RESULTS:** No serious adverse events related to the intervention were reported, and follow-up MRI showed no new MS activity in either arm. Mean nadir SpO_2_ during AIH was 84.7 ± 0.2%, and cardiovascular parameters returned to baseline within minutes of each session completion. Ankle plantarflexion torque was significantly greater following AIH than Sham (β = 8.74 N·m, p < 0.001), with mean increases of 17.5% at post-intervention and 29.2% at follow-up. Six-minute walk distance was also significantly greater after AIH (β = 10.14 m, p = 0.039). Dorsiflexion torque (p = 0.202), T25FW time (p = 0.239), and MSWS-12 score (p = 0.564) did not differ significantly between conditions. Greater elevation in heart rate during hypoxic episodes was associated with larger strength gains at follow-up (r = 0.58, p = 0.009).

**CONCLUSIONS:** Daily AIH sessions are safe and well tolerated in persons with MS across a clinically heterogeneous disability range and produce significant improvements in ankle plantarflexion strength and walking endurance. These findings support combining AIH with task-specific gait training in future trials.

## 1. Introduction

Walking impairment is among the most common and disabling consequences of multiple sclerosis (MS), affecting up to three-quarters of persons with MS at some point during their lifetime.^1,2^ Weakness, spasticity, and impaired motor coordination reduce walking speed, endurance, and independence, and walking difficulty is consistently rated by patients as the most burdensome aspect of the disease.^2,3^ Although disease-modifying therapies have reduced relapse rates, their effectiveness in addressing established motor impairments remains limited, and current rehabilitative approaches are constrained by insufficient neural activation as a result of multifocal demyelinated plaques.^4^ Approaches that augment neuroplasticity and strengthen residual neural connections are therefore needed.

Acute intermittent hypoxia (AIH) is a noninvasive means of inducing spinal neuroplasticity by delivering brief, repeated episodes of mild hypoxia alternating with room air. AIH activates carotid body chemoreceptors, triggering episodic serotonin release in the spinal cord, which initiates the synthesis of brain-derived neurotrophic factor (BDNF) and activates its high-affinity receptor, TrkB, thereby strengthening synaptic input to spinal motor neurons.^5,6^ This plasticity is expressed as a rapid increase in voluntary motor output, emerging within 60 to 90 minutes of exposure.^7^ Critically, therapeutic AIH differs fundamentally from the pathological intermittent hypoxia of obstructive sleep apnea; the low dose and episodic structure of therapeutic protocols engage plasticity-promoting pathways without inducing maladaptive cardiovascular or neurological changes.^8,9^

In humans with incomplete spinal cord injury (SCI), five consecutive days of AIH improved walking speed and endurance, with gains that persisted for weeks after treatment.^10^ Combining AIH with overground walking practice yielded greater gains in walking endurance than either intervention alone, demonstrating that AIH is most effective as a plasticity-promoting primer when paired with task-specific training.^10,11^ AIH has also increased ankle plantarflexion torque in persons with incomplete SCI after a single session^12^ and enhanced corticospinal excitability in neurologically intact individuals and in individuals with SCI.^13,14^ Across these studies, AIH was safe and well-tolerated with no serious adverse events.

Despite this evidence, the application of AIH in MS remains largely unexplored. A single AIH session improved ankle torque and cognitive processing speed in persons with relapsing-remitting MS, supporting the presence of AIH-responsive neuroplasticity in this population.^4^ Whether repeated daily AIH is safe over five consecutive days in persons with MS, and whether it produces meaningful changes in walking and somatic motor function, has not been established. This is a critical gap given that MS-related demyelination may alter the neurochemical substrates of AIH-induced plasticity, and that the safety profile of repeated AIH in this population requires prospective evaluation. The present study tested the hypothesis that five consecutive days of AIH are safe and well-tolerated in persons with MS and lead to improvements in ankle strength and walking performance compared with Sham.

## 2. Methods

### 2.1 Study design

We used a randomized, double-masked, placebo-controlled crossover study design (Figure 1). Participants were assigned to receive both AIH and Sham AIH, in a random order, with a minimum 4-week washout period between blocks to reduce carryover effects. Treatment allocation was concealed from participants and all outcome assessors throughout the study. Outcome assessments were conducted on Day 1, before the first intervention session, and on Day 5, after the final session. A follow-up assessment was conducted within the first week after Day 5. Days 2 through 4 consisted solely of intervention sessions. The Northwestern University Institutional Review Board approved this study (STU00219535), and all participants provided written informed consent prior to enrollment (ClinicalTrials.gov: NCT06276634).

**Figure 1.**
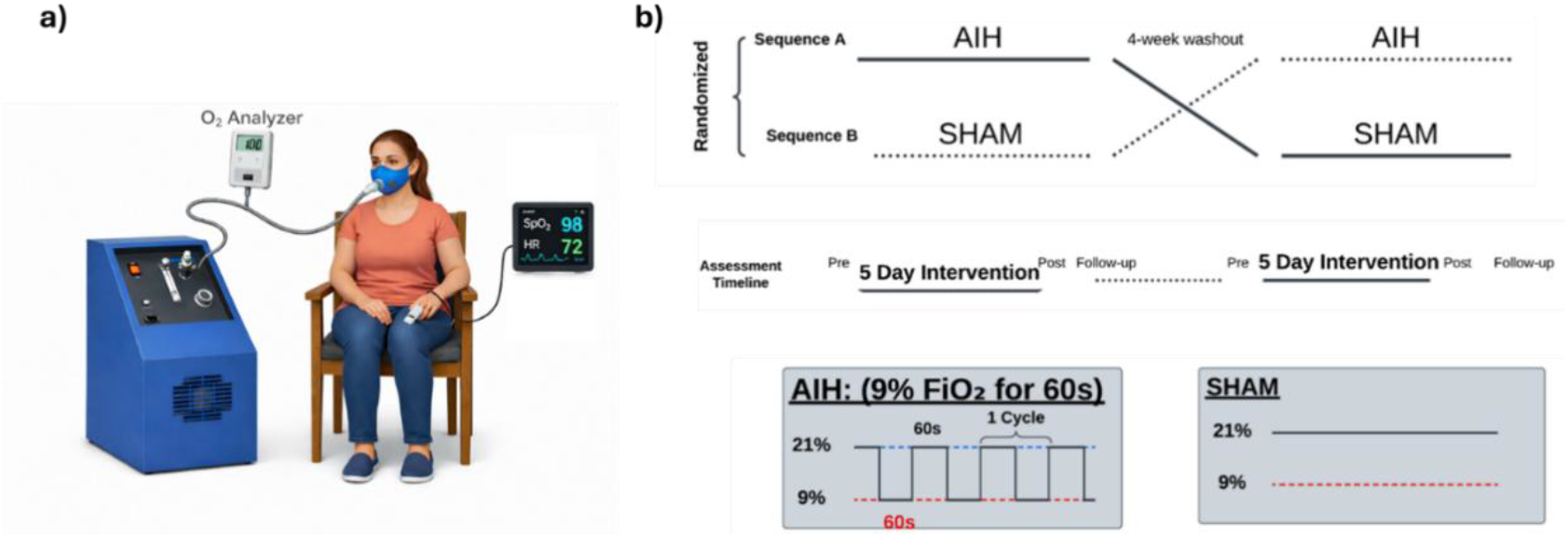
Study design and intervention protocol. (a) Participants breathed through a non-rebreathing facemask connected to a hypoxic gas generator, with SpO_2_ and heart rate continuously monitored. (b, top) Randomized, double-masked crossover design with a minimum 4-week washout separating the two intervention blocks. Each block included pre- and post-intervention assessments, as well as a follow-up visit within 1 week of the final session. (b, bottom) A single AIH session consisted of 15 hypoxic episodes (FiO_2_ = 0.09, ∼60 s, titrated to SpO_2_ 84-88%) alternating with ∼60 s normoxic intervals (FiO_2_ = 0.21). Sham sessions delivered room air (FiO_2_ = 0.21) throughout an identical timing structure.

#### 2.2 Participants

Participants were recruited from the neurology clinics of Northwestern Memorial Hospital and the Shirley Ryan AbilityLab Clinical Research Registry. Of 180 persons contacted, 25 enrolled and 23 completed both intervention blocks (Figure 2). Inclusion criteria were a confirmed diagnosis of relapsing-remitting or secondary-progressive MS, Expanded Disability Status Scale (EDSS) score between 3.0 and 6.5, at least one year relapse-free, age between 18 and 75 years, and clearance for MRI based on an institutional screening questionnaire. EDSS was assessed by a neurologist on the study team. Participants taking dalfampridine were eligible if the dose had been stable for at least two months prior to screening. We excluded persons with any active lesions on baseline MRI, uncontrolled blood pressure (systolic outside 85 to 160 mmHg or diastolic outside 55 to 110 mmHg), history of epilepsy, chronic obstructive pulmonary disease, uncontrolled sleep apnea, inability to undergo MRI, ankle spasticity greater than 3 on the Modified Ashworth Scale, diagnosis of Long COVID-19, Mini-Mental State Examination (MMSE) score of 20 or below, or confirmed pregnancy.

**Figure 2.**
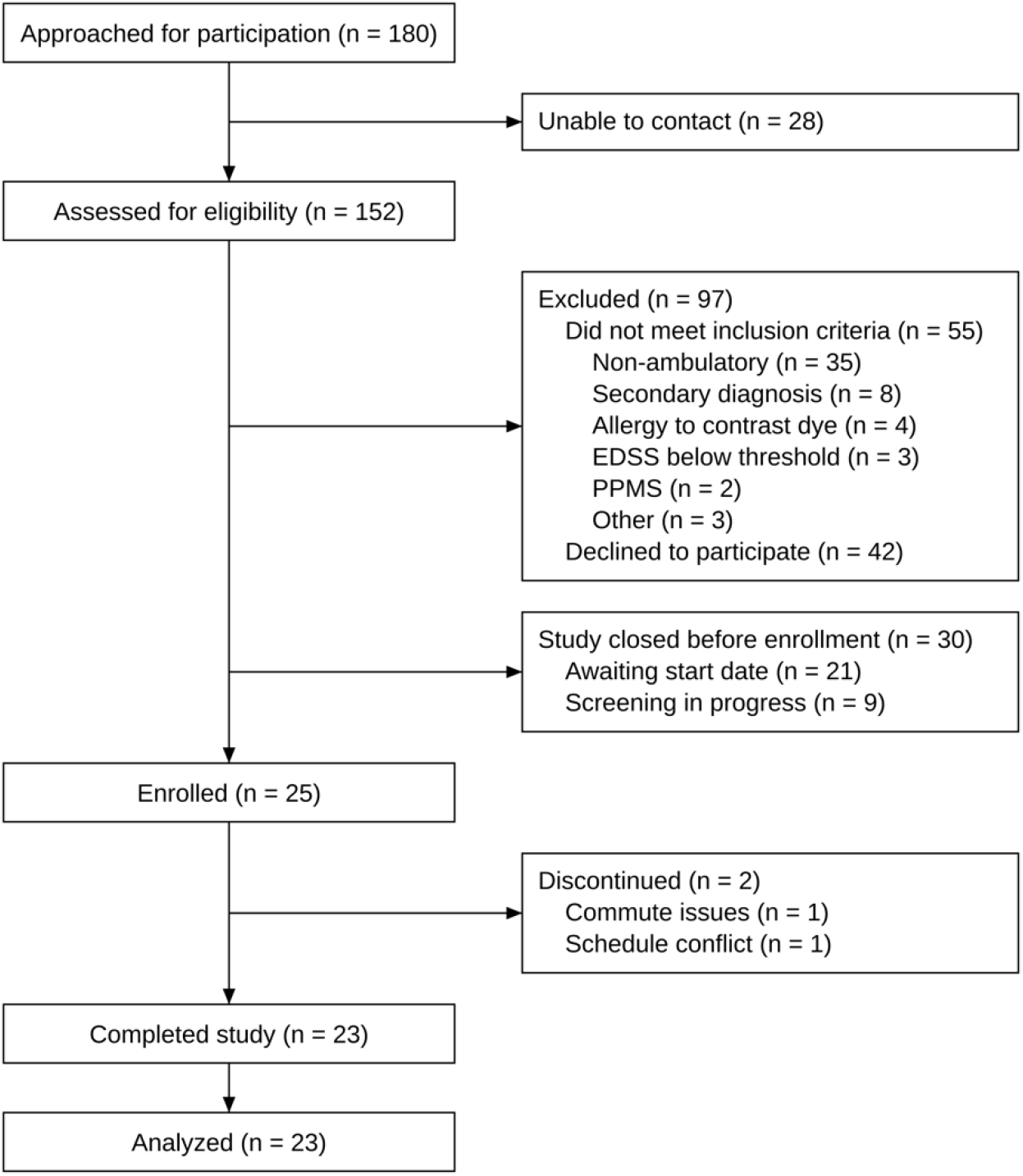
Participant flow through the study. Of 180 individuals approached, 28 could not be contacted and 152 were assessed for eligibility. Of these, 97 were excluded (55 did not meet inclusion criteria; 42 declined to participate), and 30 were still awaiting a start date (n = 21) or being screened (n = 9) when the study closed to enrollment. Twenty-five participants were enrolled, 2 discontinued (commute issues, n = 1; schedule conflict, n = 1), and 23 completed the study. EDSS, Expanded Disability Status Scale; PPMS, primary progressive multiple sclerosis.

The 23 participants who completed the study included 13 women and 10 men with a mean age of 58.8 ± 11.6 years. Fifteen had relapsing-remitting MS, and 8 had secondary-progressive MS. The mean time since diagnosis was 21.6 ± 13.6 years, and the mean EDSS score was 5.4 ± 1.2 (range 3.0 to 6.5). Participant characteristics are summarized in Table 1.

**Table 1.**
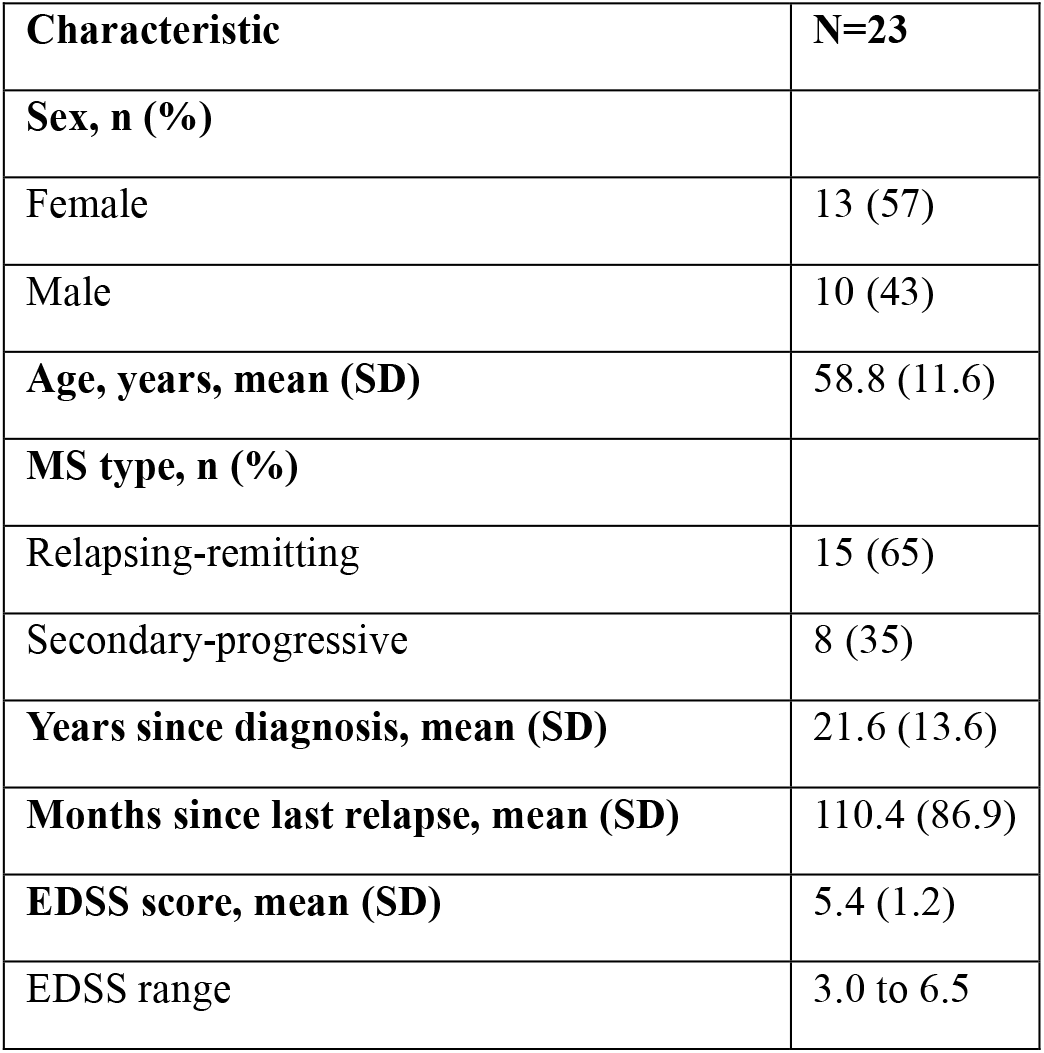
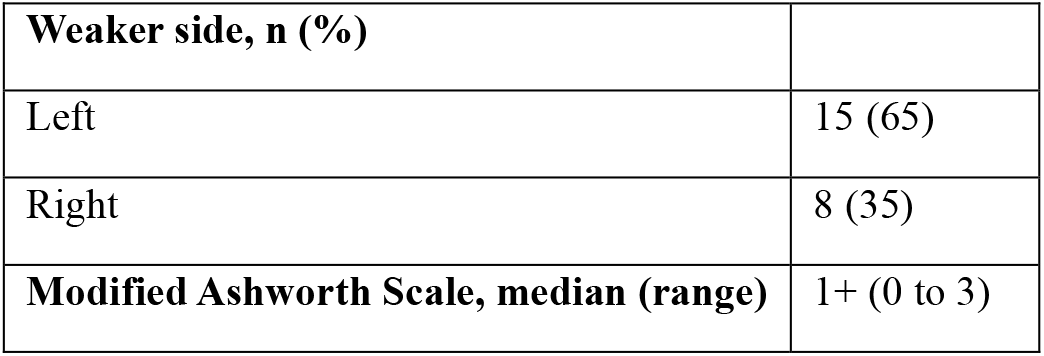
Baseline participant characteristics.

### 2.3 Intervention

#### 2.3.1 Air Delivery System

We administered AIH and Sham AIH using a Hypoxico HYP-123 hypoxic air generator (Hypoxico Inc., New York, NY), consistent with prior AIH studies in humans with neurological conditions.^10,11,15-17^ Participants were seated and fitted with a non-rebreathing face mask connected to the generator via an inlet line. An unblinded team member administered AIH or Sham AIH using controlled air delivery from the hypoxic air generator at timed intervals. When the inlet line was attached, participants inhaled air supplied directly from the generator (either hypoxic or normoxic air). When the inlet line was detached, participants breathed room air (21% oxygen) from the surrounding environment. Participants remained blinded to intervention assignment throughout.

#### 2.3.2 AIH Protocol

Each AIH session consisted of 15 episodes delivered over approximately 30 minutes. Each episode consisted of one hypoxic bout followed by one normoxic interval. During hypoxic bouts, the generator delivered air with an inspired oxygen fraction (FiO2) of 0.09.

We titrated hypoxic bout duration individually in each session to achieve a target arterial oxygen saturation (SpO_2_) of 84-88%. The first hypoxic bout of every session was fixed at 40 seconds to assess each participant’s SpO_2_ response before titration. For all subsequent bouts, the administrator adjusted bout duration in 5-second increments based on the SpO_2_ nadir achieved in the preceding bout. If SpO_2_ did not reach the target range, bout duration was extended by 5 seconds for the next episode; if SpO_2_ fell below the target range, bout duration was reduced by 5 seconds. Normoxic intervals lasted at least 60 seconds or until SpO_2_ recovered to at least 92%, whichever was longer. To facilitate consistent and reproducible timing, the subsequent hypoxic bout was initiated at the next whole-minute mark after SpO_2_ recovery.

#### 2.3.3 Sham Protocol

The Sham protocol was administered identically to AIH, including the same mask setup, inlet line attachment and detachment sequence, and session timing. The generator delivered room air (21% oxygen) throughout all bouts, producing no hypoxic stimulus. Participants were unable to distinguish AIH from Sham based on the breathing experience.

#### 2.3.4 Physiological Monitoring

We continuously monitored SpO_2_ and heart rate throughout each session using a Nonin Model 7500 pulse oximeter (Nonin Medical Inc., Plymouth, MN), with the sensor placed on the finger or ear based on signal quality. Blood pressure was measured before the start of each session and again 5 minutes after session completion using a standard automated cuff. Pre-session resting values of SpO_2_ and heart rate were recorded before mask application. Post-session values were recorded 5 minutes after session completion. Nadir SpO_2_ and the corresponding heart rate were recorded for each episode. Sessions were discontinued immediately if SpO_2_ fell below 75%, heart rate exceeded 160 bpm, systolic blood pressure exceeded 160 mmHg, or if the participant reported pain, lightheadedness, dizziness, shortness of breath, or any other discomfort.

### 2.4 Outcomes

#### 2.4.1 Safety Outcomes

Adverse events were recorded by the session administrator throughout each visit, including headaches, pain, lightheadedness, dizziness, respiratory distress, cyanosis, spasms, and autonomic dysreflexia, as reported by the participant or observed directly by the administrator. Any session discontinuations and their reasons were documented.

Clinical MRI of the brain, comprising diffusion-weighted, T2-weighted fluid-attenuated inversion recovery (FLAIR), and gadolinium-enhanced T1-weighted sequences, was acquired at baseline and repeated approximately one week after each intervention block. All scans were reviewed independently by a neuroradiologist (A.J.N.) blinded to intervention assignment for new or enlarging T2/FLAIR lesions, gadolinium-enhancing lesions, or diffusion abnormalities indicative of subclinical (silent) MS activity.

Physiological tolerability was characterized from the monitoring data described in Section 2.3.4, namely nadir SpO_2_ and the corresponding heart rate for each hypoxic episode, resting SpO_2_ and heart rate before and after each session, and pre- and post-session blood pressure.

#### 2.4.2 Somatic Motor Outcomes

##### Ankle Torque

Maximum voluntary isometric plantarflexion and dorsiflexion torques of the weaker limb were measured with a six degree-of-freedom load cell attached to a Biodex System 4 dynamometer (Biodex Medical Systems, Shirley, NY, USA). Participants were secured in the dynamometer and instructed to push (plantarflexion) or pull (dorsiflexion) against the footplate for 3 s. Three attempts were performed in each direction at each time point, separated by rest intervals of at least 30 s, and the highest peak torque across the three attempts was taken as the maximum voluntary contraction (MVC). Torque and EMG signals were digitized with a Power1401 interface and recorded in Spike2 software (Cambridge Electronic Design, Cambridge, UK).

##### Electromyography

Surface EMG was recorded during plantarflexion and dorsiflexion attempts from the medial gastrocnemius, soleus, tibialis anterior, and quadriceps muscles with single differential electrodes placed according to the Surface ElectroMyoGraphy for the Non-Invasive Assessment of Muscles (SENIAM) guidelines. Signals were acquired with a Bagnoli EMG system (Delsys Inc., Natick, MA, USA), amplified 1000 times, band-pass filtered (20-450 Hz), sampled at 2000 Hz, and synchronized with the torque signal. EMG data were rectified, smoothed, and integrated offline in Spike2. Peak EMG activity for each attempt was computed over a 500 ms window extending from 100 ms before to 400 ms after the maximum absolute value of the torque signal, and peak values were averaged across the three attempts for each muscle. EMG activation was expressed as the percent increase in peak EMG relative to resting EMG for the soleus during plantarflexion and for the tibialis anterior during dorsiflexion.

#### 2.4.3 Walking Outcomes

##### Six-Minute Walk Test

Walking endurance was quantified using the 6MWT.^18^ Participants walked along a standardized indoor course approximately 60 meters long at a maximal sustainable speed for 6 minutes. No physical assistance or facilitation was permitted. Participants used their customary assistive device throughout all time points. The total distance covered in 6 minutes was recorded in meters.

##### Timed 25-Foot Walk Test

Walking speed was quantified using the T25FW, a component of the MS Functional Composite.^19^ Participants were instructed to walk a clearly marked 25-foot course at maximum safe effort. Two trials were performed at each time point, and the mean time across both trials was used for analysis. Assistive devices were permitted and kept consistent across visits.

##### 12-Item MS Walking Scal

Self-reported walking limitation was assessed using the MSWS-12.^20^ The scale comprises 12 items, each scored on a 5-point scale, summed to a total score ranging from 12 to 60; higher scores indicate greater walking limitation.

### 2.5 Data Analysis

#### 2.5.1 Primary Analysis

Prior to primary analyses, potential carryover effects were assessed separately for each outcome using independent-samples t-tests, with the sum of each participant’s outcome values across both intervention periods as the dependent variable and treatment sequence as the grouping factor. Where no significant carryover was detected, the full crossover model was applied.

Treatment effects were estimated using analysis of covariance (ANCOVA) models fitted separately for each outcome. Each model included the following fixed effects: treatment condition (AIH vs. Sham), treatment sequence, intervention period, the difference between each participant’s two period baselines for the outcome (period 1 minus period 2), the interaction of that baseline difference with period, and assessment timepoint (post-intervention vs. follow-up). All models were adjusted for age, sex, MS type (relapsing-remitting vs. secondary-progressive), years since MS diagnosis, EDSS score, and baseline ankle plantarflexion torque (the pre-intervention value from the first intervention block); the baseline torque covariate was omitted from the plantarflexion torque model. Participants were modeled as a random effect with an unstructured covariance matrix to account for the repeated-measures structure of the crossover design. Where the treatment-by-timepoint interaction was not statistically significant, the interaction term was removed, and a main effect of treatment was tested. For EMG outcomes, sessions in which the averaged peak EMG of a muscle did not exceed its resting EMG were treated as invalid recordings for that muscle and were excluded from analysis; otherwise, the EMG analyses used the same participants and sessions as the torque analyses. All tests were two-sided with a significance threshold of p < 0.05. Analyses were performed in R^21^ and Python (Python Software Foundation, Wilmington, DE).

#### 2.5.2 Exploratory Associations

To characterize individual responsiveness to AIH, Pearson correlations were computed between two physiological variables and the AIH-induced change in strength. Strength change was defined as the mean percent change from pre-intervention to follow-up in ankle plantarflexion and dorsiflexion torque during the AIH condition. The variables were the mean percent change in SpO_2_ and in heart rate during hypoxic episodes (nadir SpO_2_ and the corresponding heart rate relative to normoxic intervals), averaged across all 15 episodes per session over the 5 AIH sessions, to assess whether the degree of physiological response to hypoxia was associated with strength response. Pearson correlation coefficients (r) and associated p-values are reported. A significance threshold of p < 0.05 was applied.

## 3. Results

Of 180 individuals approached for participation, 28 could not be contacted and 152 were assessed for eligibility (Figure 2). Ninety-seven were excluded: 55 did not meet inclusion criteria (35 unable to walk, 8 with a secondary diagnosis, 4 with contrast dye allergy, 3 with EDSS below the inclusion threshold, 2 with primary progressive MS, and 3 for other reasons), and 42 declined to participate. A further 30 were awaiting a start date (n = 21) or still being screened (n = 9) when the study closed to enrollment. Twenty-five participants provided written informed consent and were randomized; 2 withdrew before completing both intervention blocks (1 due to commute issues, 1 due to scheduling conflicts) and were not included in any analyses. Twenty-three participants completed both intervention blocks. Walking outcomes (6MWT, T25FW, MSWS-12) were obtained from all 23 participants, and Biodex torque and EMG assessments from 20; the remaining 3 participants did not complete the Biodex assessments. Because baseline plantarflexion torque was a covariate in the dorsiflexion and walking models and the plantarflexion model required Biodex data, each ANCOVA included these 20 participants (plantarflexion and dorsiflexion torque, 78 observations each; 6MWT and T25FW, 80 observations each; MSWS-12, 79 observations). After excluding invalid EMG recordings, the soleus model included 17 participants (63 observations), and the tibialis anterior model included 18 participants (68 observations). Participant demographic and clinical characteristics are summarized in Table 1.

### 3.1 Safety and tolerability

#### 3.1.1 Adverse events

No serious adverse events related to the intervention were recorded during either the AIH or Sham intervention blocks. Two expected adverse events occurred in one participant during the AIH block. Immediately after the first session, the participant reported mild, transient dizziness lasting about 60 s, similar to dizziness previously experienced with optic neuritis; vital signs remained stable. Nine minutes into the second session, the participant reported feeling slightly off, with a sensation of eye misalignment; blood pressure was 101/61 mmHg, and symptoms resolved within 17 minutes. This session was terminated, and the participant completed the remaining sessions without any adverse events after physician review. Two fall-related incidents occurred for one participant during assessment visits: one during a wheelchair transfer and one during the 6MWT. In both cases, the assessor lowered the participant to the floor, and no injury or discomfort was reported. Two serious adverse events were recorded, neither of which was attributable to the intervention. One participant fell at home before the intervention period and was later diagnosed with a lacerated right kidney, which resolved without surgical intervention; the study team judged the event unrelated to the study. Another participant fell twice during EDSS assessment without injury and reported that falls were a daily occurrence; a follow-up call two days later confirmed no injury. The event was reported to the IRB, and fall-risk language was added to the informed consent form and protocol. No adverse events resulted in withdrawal from the study. All participants completed follow-up MRI after both the AIH and Sham intervention blocks, and no new or enlarging T2/FLAIR lesions, gadolinium-enhancing lesions, or diffusion abnormalities were identified in any participant in either arm.

#### 3.1.2 Physiological response during AIH

Cardiovascular responses to AIH and Sham are illustrated in Figure 3 and summarized across all participants and sessions. During AIH, mean nadir SpO_2_ during hypoxic bouts was 84.7 ± 0.2% (standard error of the mean, SEM), consistent with the target range of 84-88%, compared to 96.2 ± 0.2% during the corresponding bouts in the Sham condition (t = -40.83, p < 0.001, Figure 3, top). During normoxic intervals, mean SpO_2_ was 95.4 ± 0.3% for AIH and 96.6 ± 0.3% for Sham, confirming adequate recovery between hypoxic episodes. SpO_2_ did not fall below 75% in any participant at any point during AIH sessions. During Sham, SpO_2_ remained stable throughout all bouts (96.2 ± 0.2%), with no significant difference between normoxic intervals and condition bouts.

**Figure 3.**
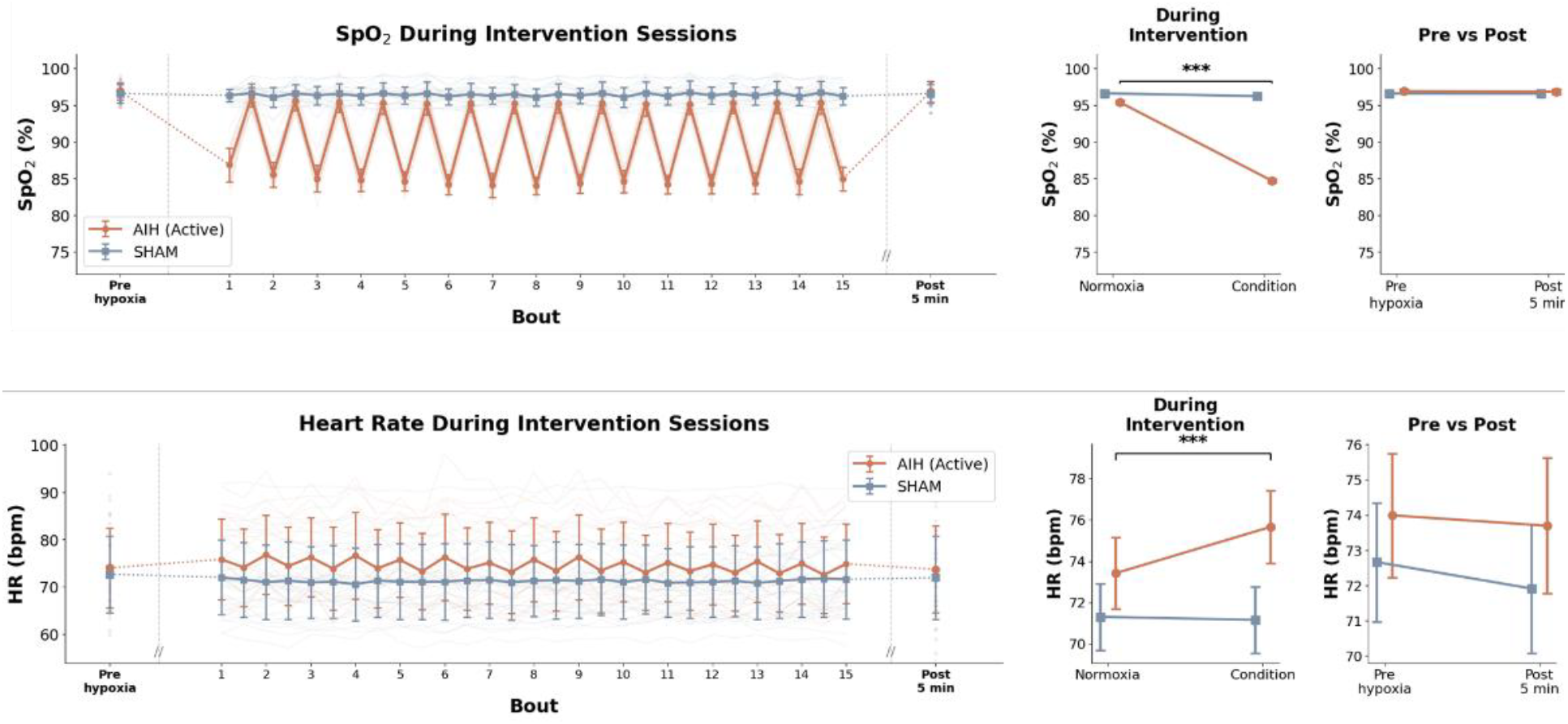
Cardiovascular responses during AIH and Sham intervention sessions. SpO_2_ (%, top row) and heart rate (bpm, bottom row) across all 15 bouts for AIH (orange) and Sham (blue), averaged across all participants and sessions. Large markers indicate group means ± SEM; faint lines represent individual participant data. Dotted lines connect pre-session and post-session values to the main session data. The middle panels show mean ± SEM during normoxic intervals and condition bouts. The right panels show pre-session and post-session resting values. *** p < 0.001.

Heart rate increased significantly during hypoxic bouts of AIH compared with normoxic intervals (75.7 ± 1.8 vs 73.4 ± 1.7 bpm; t = 3.81, p < 0.001; Figure 3, bottom), consistent with a chemoreflex-mediated response to hypoxia. Heart rate during Sham condition bouts remained stable and comparable to normoxic intervals (71.2 ± 1.6 vs 71.3 ± 1.6 bpm).

Resting SpO_2_ and heart rate were comparable before and after intervention sessions in both conditions. Pre-session SpO_2_ was 96.9 ± 0.3% for AIH and 96.6 ± 0.3% for Sham; post-session values were 96.8 ± 0.3% and 96.6 ± 0.3%, respectively, with no significant change in either condition (AIH: t = 0.38, p = 0.707; Sham: t = 0.10, p = 0.920). Pre-session heart rate was 74.0 ± 1.8 bpm for AIH and 72.7 ± 1.7 bpm for Sham; post-session values were 73.7 ± 1.9 and 71.9 ± 1.8 bpm, respectively, with no significant change in either condition (AIH: t = 0.71, p = 0.487; Sham: t = 1.71, p = 0.101). Blood pressure returned to pre-session baseline values within 5 to 10 minutes of session completion in all participants.

### 3.2 Somatic motor outcomes

Ankle plantarflexion and dorsiflexion MVC torques are shown in Figure 4. Of 23 participants, 20 completed Biodex assessments and 18 completed all six assessment visits; the remaining participants were unable to complete one or more assessments due to scheduling and clinical constraints. Percent changes were calculated as the mean of individual participant changes relative to the pre-intervention baseline (Figure 4).

**Figure 4.**
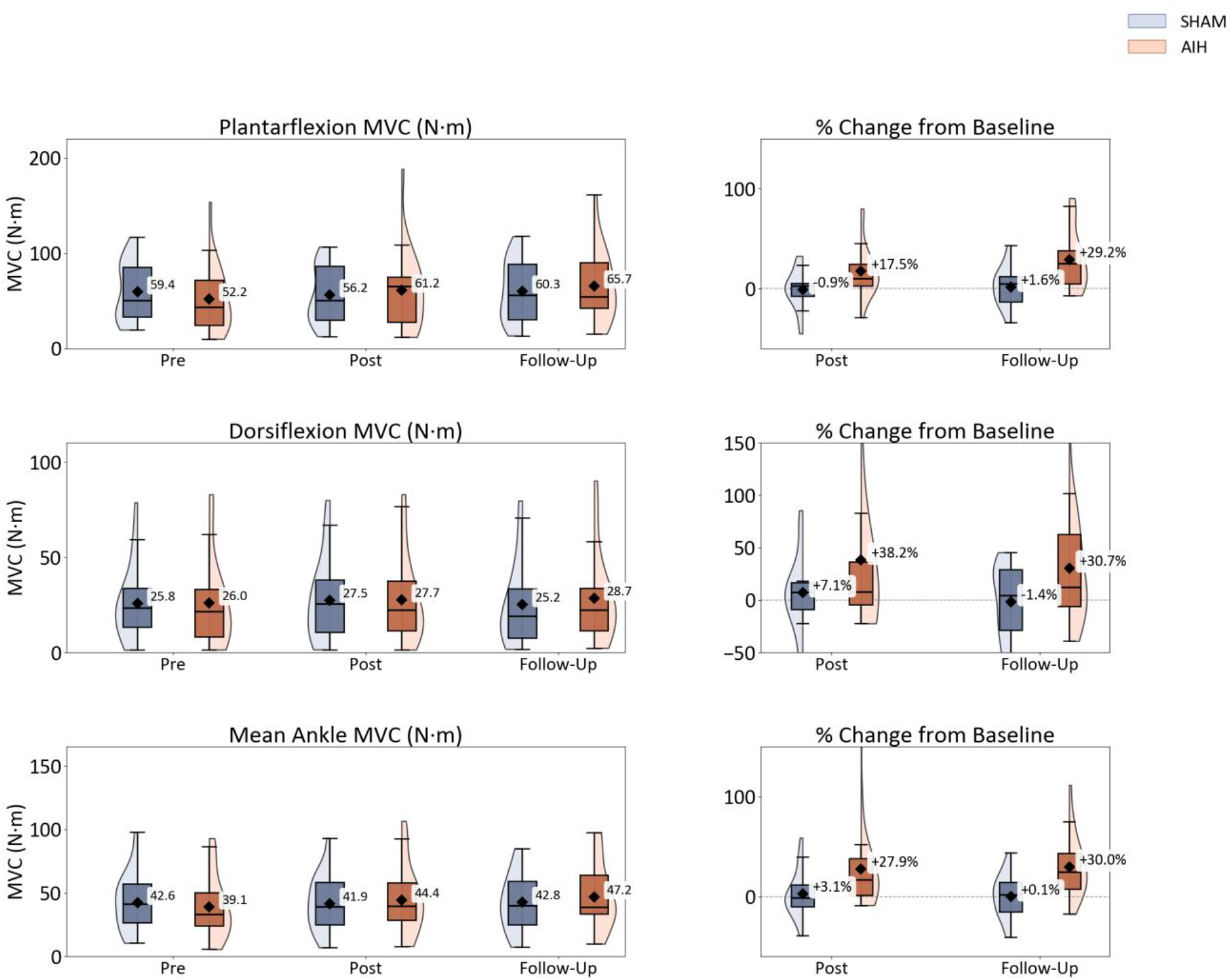
Ankle plantarflexion torque, dorsiflexion torque, and mean ankle torque (the average of plantarflexion and dorsiflexion) at pre-intervention, post-intervention, and follow-up for AIH (orange) and Sham (blue). The left panels show absolute maximum voluntary contraction (MVC) torque values, and the right panels show the mean individual percent change from the pre-intervention baseline. Diamonds indicate group means, boxes indicate the interquartile range, and shaded areas show the distribution of individual values.

Plantarflexion torque was significantly greater following AIH than Sham (β = 8.74 N·m, SE = 2.40, p < 0.001). Mean plantarflexion torque increased from 52.2 ± 36.2 N·m at pre-intervention to 61.2 ± 42.5 N·m at post-intervention and 65.7 ± 38.2 N·m at follow-up during AIH (+17.5% and +29.2% relative to pre-intervention), compared with 59.4 ± 31.1 N·m, 56.2 ± 29.7 N·m, and 60.3 ± 32.3 N·m during Sham (-0.9% and +1.6% respectively). A significant period effect (p = 0.015) and baseline-by-period interaction (p < 0.001) were accounted for in the model.

Dorsiflexion torque did not differ significantly between AIH and Sham (β = 1.75 N·m, SE = 1.35, p = 0.202). Mean dorsiflexion torque was 26.0 ± 24.7 N·m, 27.7 ± 23.2 N·m, and 28.7 ± 24.3 N·m at pre-intervention, post-intervention, and follow-up during AIH (+38.2% and +30.7% relative to pre-intervention), compared with 25.8 ± 20.1 N·m, 27.5 ± 22.4 N·m, and 25.2 ± 22.4 N·m during Sham (+7.1% and -1.4% respectively). The adjusted treatment effect did not reach statistical significance. The baseline-by-period interaction was significant (p = 0.027). Mean ankle torque (the average of plantarflexion and dorsiflexion), which served as the strength outcome in the exploratory analyses, increased by 27.9% at post-intervention and 30.0% at follow-up during AIH, compared with 3.1% and 0.1% during Sham (Figure 4, bottom).

#### Muscle activation

EMG activation, expressed as the percent increase in peak EMG relative to resting EMG, was analyzed in the same participants and sessions as the torque outcomes, after excluding invalid recordings (soleus: 7 of 118 sessions; tibialis anterior: 8 of 118 sessions). Soleus activation during plantarflexion did not differ between AIH and Sham (β = 151 percentage points, SE = 105, 95% CI: -60 to 363, p = 0.16; 17 participants, 63 observations), although the direction of the difference favored AIH, and peak soleus EMG was 41% higher after AIH than after Sham (95% CI: -2% to 103%, p = 0.064). Tibialis anterior activation during dorsiflexion did not differ between conditions (β = 78 percentage points, SE = 346, 95% CI: -619 to 776, p = 0.82; 18 participants, 68 observations). No treatment-by-timepoint interaction (p = 0.30 and 0.71) or carryover effect (p ≥ 0.30) was detected for either muscle.

### 3.3 Walking Outcomes

Walking outcomes for all three measures are presented in Figure 5. Percent changes were calculated as the mean of individual participant changes relative to the pre-intervention baseline; group means of the absolute values are also reported. Descriptive values are based on all 23 participants; treatment effects were estimated in the 20 participants with baseline torque data.

**Figure 5.**
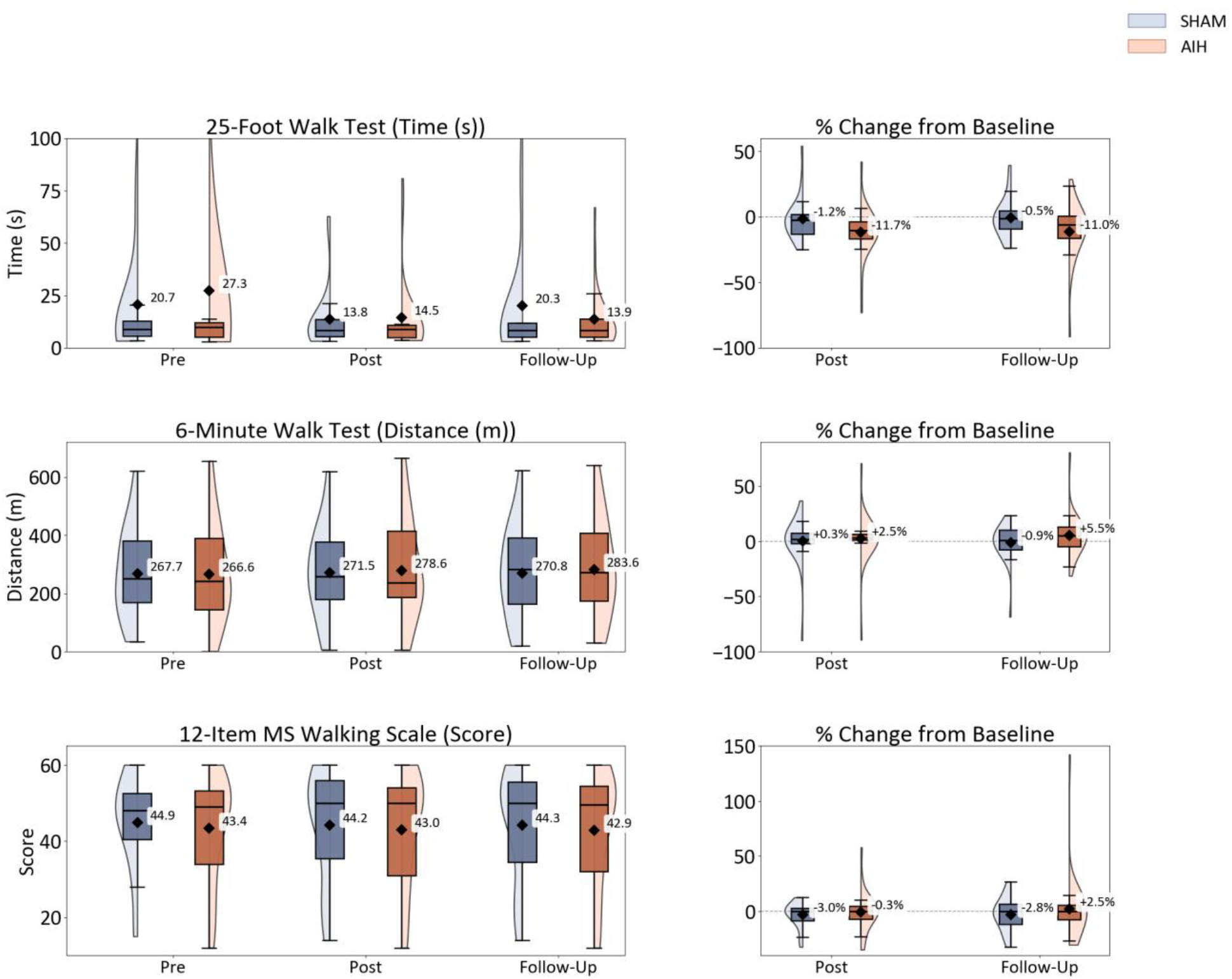
Timed 25-Foot Walk Test time (s), Six-Minute Walk Test distance (m), and MSWS-12 score at pre-intervention, post-intervention, and follow-up for AIH (orange) and Sham (blue). The left panels show absolute values, and the right panels show the mean individual percent change from the pre-intervention baseline. Diamonds indicate group means, and boxes indicate the interquartile range. Lower T25FW times indicate faster walking.

#### Six-Minute Walk Test

Six-minute walk distance was significantly greater following AIH than Sham (β = 10.14 m, SE = 4.81, p = 0.039). Following AIH, the mean 6MWT distance was 266.6 ± 169.2 m at pre-intervention, 278.6 ± 175.2 m at post-intervention, and 283.6 ± 174.3 m at follow-up, corresponding to mean individual percent changes of +2.5% and +5.5%, respectively, relative to pre-intervention. Following Sham, the mean 6MWT distance was 267.7 ± 157.9 m at pre-intervention, 271.5 ± 156.1 m at post-intervention, and 270.8 ± 161.4 m at follow-up, corresponding to mean individual percent changes of +0.3% and -0.9%, respectively. A significant sequence effect (p = 0.004) and a significant baseline-by-period interaction (p < 0.001) were observed; these effects were accounted for in the ANCOVA model.

#### Timed 25-Foot Walk Test

T25FW time did not differ significantly between AIH and Sham (β = -0.31 s, SE = 0.26, p = 0.239). Following AIH, mean T25FW time was 27.3 ± 60.9 s at pre-intervention, 14.5 ± 19.7 s at post-intervention, and 13.9 ± 15.1 s at follow-up, corresponding to mean individual percent changes of -11.7% and -11.0%, respectively, relative to pre-intervention. Following Sham, mean T25FW time was 20.7 ± 34.7 s at pre-intervention, 13.8 ± 16.1 s at post-intervention, and 20.3 ± 31.2 s at follow-up, corresponding to mean individual percent changes of -1.2% and -0.5%, respectively. Although the mean T25FW time decreased in the AIH condition, the adjusted treatment effect did not reach statistical significance. Baseline plantarflexion torque (p = 0.025) and the baseline difference (p = 0.027) were significant covariates in this model.

#### 12-Item MS Walking Scale

MSWS-12 score did not differ significantly between AIH and Sham (β = 0.69, SE = 1.19, p = 0.564). Mean MSWS-12 score in the AIH condition was 43.4 ± 14.3 at pre-intervention, 43.0 ± 15.2 at post-intervention, and 42.9 ± 15.6 at follow-up, corresponding to mean individual percent changes of -0.3% and +2.5%, respectively, relative to pre-intervention. In the Sham condition, the mean MSWS-12 score was 44.9 ± 12.8 at pre-intervention, 44.2 ± 15.0 at post-intervention, and 44.3 ± 15.3 at follow-up, corresponding to mean individual percent changes of -3.0% and -2.8%, respectively. The baseline-by-period interaction was significant (p < 0.001) and was accounted for in the model.

Across models, EDSS score was a significant covariate for plantarflexion torque (p = 0.015), dorsiflexion torque (p = 0.022), 6MWT distance (p = 0.004), and MSWS-12 score (p = 0.004), with higher EDSS associated with lower torque, shorter 6MWT distance, and higher MSWS-12 score.

### 3.4 Exploratory associations with strength change

Associations between participant-level variables and AIH-induced strength change (the mean percent change in ankle plantarflexion and dorsiflexion torque from pre-intervention to follow-up in the AIH condition) were examined in the 19 participants with follow-up torque data. A greater mean rise in heart rate during hypoxic episodes was associated with a larger strength gain (r = 0.58, p = 0.009), whereas the mean fall in SpO_2_ during hypoxic episodes was not (r = -0.17, p = 0.485).

## 4. Discussion

This randomized, double-masked, placebo-controlled crossover trial examined the safety and preliminary effects of five consecutive days of AIH in persons with MS. Three key findings emerged from this study. First, daily AIH was safe and well tolerated across all sessions; physiological responses remained within acceptable limits, and no intervention-related serious adverse events were recorded. Second, AIH produced significant improvements in ankle plantarflexion torque and six-minute walk distance compared to Sham, whereas dorsiflexion torque, T25FW time, and MSWS-12 score did not differ significantly between conditions. Third, a greater rise in heart rate during AIH hypoxic episodes was associated with greater gains in ankle strength at follow-up, suggesting that the magnitude of autonomic chemoreflex engagement may influence individual responsiveness to AIH. Together, these preliminary findings support the feasibility and safety of repeated daily AIH and suggest potential motor benefits in persons with MS.

### 4.1 Safety of Repeated AIH in MS

A primary objective of this study was to establish the safety and tolerability of five consecutive daily AIH sessions in persons with MS. AIH was well tolerated across all participants and sessions. Mean nadir SpO_2_ during hypoxic episodes was 84.7 ± 0.2%, consistent with the target range of 84-88% and comparable to nadir values reported in prior human AIH studies in SCI.^10,11^ SpO_2_ and heart rate returned to pre-session baseline values within 5 minutes of session completion in both conditions, and blood pressure normalized within 5 to 10 minutes, indicating no sustained cardiovascular perturbation. No participant withdrew due to an adverse event.

These findings extend the safety profile of AIH established in persons with SCI or stroke to persons with MS. Prior work with a single AIH session in ten individuals with relapsing-remitting MS reported no adverse events and well-maintained cardiovascular responses.^4^ The current study demonstrates that this safety profile is maintained across multiple sessions in a larger and more disability-heterogeneous sample, including individuals with secondary-progressive MS and EDSS scores up to 6.5. Independent, blinded neuroradiological review of contrast-enhanced MRI acquired after each intervention block revealed no evidence of subclinical MS activity in either arm, indicating that repeated exposure to mild hypoxia did not provoke new inflammatory lesions during the study period.

### 4.2 Effects on Somatic motor output at the ankle

Five days of AIH increased ankle plantarflexion torque relative to Sham, and the gain was maintained at follow-up. These findings are consistent with prior evidence that AIH augments somatic motor output through serotonin- and BDNF-dependent mechanisms that strengthen synaptic input to spinal motor nuclei.^5,22^ Plantarflexor muscles are tonically active, slow-twitch-dominant antigravity muscles^23^ whose motor pools are enriched in low-threshold motor neurons with large persistent inward currents that depend on brainstem serotonergic input,^24,25^ making them a likely target of AIH-induced plasticity. A single AIH session previously increased ankle torque in persons with relapsing-remitting MS^4^ and in persons with incomplete SCI.^12^ The current findings demonstrate that repeating AIH over five consecutive days yields gains that persist through follow-up, suggesting cumulative or consolidating plasticity effects beyond a single session.

Dorsiflexion torque did not differ between conditions, despite a numerical trend toward improvement after AIH. The absence of a significant effect on dorsiflexion contrasts with the significant plantarflexion response. It may reflect differences in motor neuron pool excitability, corticospinal drive, or serotonergic terminal density between the muscle groups. Dorsiflexors are phasically active muscles predominantly involved in swing-phase foot clearance, and their motor neuron pools may be less amenable to tonic facilitation induced by AIH than those of the plantarflexors. Alternatively, the high within-group variability in dorsiflexion torque may have reduced statistical power to detect a treatment effect. Future trials with larger samples should examine whether the differential response between plantarflexors and dorsiflexors is a consistent feature of AIH in MS.

Surface EMG did not mirror the torque findings: soleus activation was numerically higher after AIH than after Sham, but not significantly so, and tibialis anterior activation did not differ. Surface EMG is an insensitive index of motoneuron output during maximal contractions because of amplitude cancellation^26,27^; the peak-to-rest ratio varies with resting EMG and electrode placement across sessions, and the sample was small and variable. AIH may also change motor unit discharge timing rather than surface amplitude. Motor evoked potentials or H-reflexes would better identify the neural substrate of the strength gains.

### 4.3 Effects on Walking Outcomes

Six-minute walk distance was significantly greater following AIH than Sham, with mean distance increasing by 2.5% at post-intervention and 5.5% at follow-up in the AIH condition. This improvement in walking endurance in the absence of a significant change in T25FW time suggests that AIH may preferentially enhance sustained motor output capacity rather than peak locomotor speed over short distances. This pattern is consistent with the mechanistic role of plantarflexors as the primary propulsive generators during walking; improved plantarflexion torque may directly support greater walking endurance by maintaining forward propulsion across repeated gait cycles. A similar dissociation between endurance and speed outcomes was observed in SCI, where combined AIH and walking practice produced greater gains on the 6MWT than on speed-based measures.^10,11^

T25FW time and MSWS-12 score did not differ significantly between conditions. The absence of a T25FW effect is consistent with the hypothesis that short-burst maximal walking speed is less sensitive to the motor facilitation produced by AIH alone, without concurrent task-specific walking training. The T25FW data were also highly variable. Mean T25FW time in the AIH condition decreased by nearly half from pre-intervention to post-intervention, but this change was driven by a few participants with very slow baseline walking times, as reflected by a mean individual change of about 12% and a pre-intervention standard deviation that exceeded the mean. This variability limited the statistical power to detect a treatment effect on walking speed despite the large change in the group mean. The lack of change in MSWS-12 despite objective improvements in plantarflexion torque and 6MWT distance is consistent with the temporal dissociation commonly observed between performance-based gains and self-reported walking limitations. It may reflect the relatively short intervention window and the influence of habitual activity patterns on subjective perceptions of disability.

A significant sequence effect was observed on the 6MWT. Participants randomized to Sham first were less disabled and walked farther at baseline, but after adjustment for EDSS and baseline strength, estimated distances were greater in the AIH-first (AS) sequence (β = 123 m). This asymmetry may reflect residual AIH effects persisting across the washout period when AIH was administered first, or a chance imbalance between the small sequence groups. The ANCOVA model adjusted for sequence and the baseline-by-period interaction, but the origin of this asymmetry warrants investigation in larger trials with longer washout periods.

### 4.4 Cardiorespiratory Response as a Predictor of Strength Gain

A greater rise in heart rate during AIH hypoxic episodes was associated with greater ankle strength gains at follow-up, whereas the nadir SpO_2_ drop was not. Heart rate increases during hypoxia reflect sympathetic activation driven by the carotid body chemoreflex. The same afferent pathway triggers episodic serotonin release in the spinal cord and initiates the plasticity cascade underlying AIH-induced motor facilitation.^6^ These findings suggest that participants who mounted a stronger autonomic chemoreflex response to hypoxia may have experienced greater serotonergic activation of spinal motor circuits, leading to more robust plasticity. This is consistent with evidence in humans that the magnitude of the hypoxic ventilatory response predicts the magnitude of intermittent hypoxia-induced respiratory plasticity.^28^ The absence of a significant relationship with nadir SpO_2_ is notable and suggests that the degree of oxygen desaturation alone does not determine the magnitude of motor response; the afferent neural response to hypoxia, reflected in heart rate, may be a more sensitive indicator of AIH dose at the level of the nervous system. These findings have potential implications for individualizing AIH dosing in future trials.

### 4.5 Clinical Implications

These preliminary findings suggest that five consecutive days of AIH may represent a feasible, safe, and potentially effective priming intervention for persons with MS. The improvement in plantarflexion torque and 6MWT distance, in the absence of any concurrent walking training, indicates that AIH alone is sufficient to produce objective motor gains in this population over a short intervention window. This is clinically relevant because AIH requires no active participation beyond breathing and places minimal physical demand on participants, making it accessible to individuals across a wide range of disability levels. The current sample spanned EDSS scores of 3.0 to 6.5 and included both relapsing-remitting and secondary-progressive subtypes, supporting the generalizability of the safety findings. Future work combining AIH with task-specific walking training, as demonstrated in SCI,^10,11^ may yield greater functional gains than AIH alone in persons with MS.

EDSS score was a significant predictor of plantarflexion torque, dorsiflexion torque, six-minute walk distance, and MSWS-12 score across ANCOVA models, with higher disability associated with lower torque and walking distance and higher self-reported walking limitation, consistent with the expected relationship between greater neurological disability and poorer motor and walking performance. Notably, this effect reflects baseline performance differences rather than differential treatment response, as EDSS was included as a covariate in all models. Whether baseline disability level moderates the magnitude of AIH-induced motor gains remains an important question for future work with adequate power to examine responder subgroups.

### 4.6 Limitations and Future Work

Several limitations should be acknowledged. The sample size, while adequate for a crossover pilot trial, limits power for secondary outcomes and subgroup analyses. The enrolled cohort was clinically heterogeneous, spanning EDSS scores from 3.0 to 6.5 and including both relapsing-remitting and secondary-progressive subtypes, likely contributing to variability in individual responses. Covariate analyses confirmed a significant association between EDSS and baseline performance across multiple outcomes. The significant sequence and period effects observed for several outcomes also suggest that full washout may not have been uniformly achieved within the 4-week interval, and future trials should consider longer washout periods or parallel-group designs.

These limitations point directly to three lines of future investigation. First, the present protocol delivered AIH in isolation, without concurrent task-specific training. The modest magnitude of walking gains observed here is consistent with this design choice and supports pairing AIH with targeted gait training as a logical next step. In persons with incomplete SCI, combining daily AIH with overground walking practice produced substantially larger gains in walking endurance than AIH alone, consistent with the principle that AIH acts as a plasticity-promoting primer that requires task-specific activity to consolidate functional benefit.^10,11^ Whether a comparable synergy can be elicited in persons with MS is the question we intend to address in our next trial.

Second, and related to the heterogeneity noted above, the AIH dose in the current protocol was defined by a fixed target FiO_2_ (0.09) with bout duration titrated to a group-level SpO_2_ target of 84-88%. This approach standardizes the stimulus at the airway. However, it does not account for between-participant differences in chemoreflex sensitivity or downstream serotonergic engagement, which may explain part of the variability in motor response. Our exploratory finding that a rise in heart rate during hypoxic episodes was associated with strength gains suggests that the physiological response, rather than inspired oxygen alone, may be a more sensitive marker of an effective dose. Future work should examine individualized dosing strategies titrated to participant-specific responses. It should establish dose-response relationships for the number of sessions, episode duration, and hypoxic severity in persons with MS.

Third, the present protocol delivered poikilocapnic hypoxia, in which arterial CO_2_ is allowed to fall passively as ventilation increases during hypoxic episodes. In awake humans, ventilatory long-term facilitation is expressed only when CO_2_ is held at or above resting levels during intermittent hypoxia,^29-31^ and recent evidence suggests that maintaining CO_2_ during hypoxia (hypercapnic-hypoxic AIH) may augment the magnitude of motor facilitation by sustaining cerebral perfusion and amplifying chemoreflex drive to spinal motor circuits.^32,33^ Direct comparison of poikilocapnic and hypercapnic protocols, with continuous end-tidal CO_2_ monitoring, is a natural extension of the dose-optimization work described above and is warranted to determine whether controlled CO_2_ delivery enhances therapeutic efficacy in persons with MS.

## 5. Conclusions

Five consecutive days of AIH were safe and well tolerated in persons with relapsing-remitting and secondary-progressive MS across a broad range of disability, with no serious intervention-related adverse events and no evidence of new MS activity on blinded MRI review. AIH alone increased ankle plantarflexion strength and six-minute walk distance relative to Sham, and the magnitude of the heart rate response during hypoxic episodes was associated with the size of the strength gain. These findings establish the safety and feasibility of repeated AIH in MS and provide the foundation for trials that pair AIH with task-specific gait training, the combination that has produced the largest functional benefits in spinal cord injury.

## Data Availability

All data produced in the present study are available upon reasonable request to the authors

## Acknowledgments

The authors thank the study participants for their time and commitment, and the neurology clinics of Northwestern Memorial Hospital and the Shirley Ryan AbilityLab Clinical Research Registry for assistance with recruitment.

## Author contributions

**A.N**.: Methodology; Investigation; Formal analysis; Data curation; Visualization; Writing (original draft); Writing (review and editing).

**R.K**.: Investigation (participant screening and recruitment, data collection); Project administration; Writing (review and editing).

**A.B**.: Investigation (intervention delivery, scheduling); Project administration (regulatory and data management); Writing (review and editing).

**L.O**.: Investigation (outcome assessments); Writing (review and editing).

**H.R.J**.: Investigation (neuroimaging acquisition and analysis); Writing (review and editing).

**C.Z**.: Investigation; Writing (review and editing).

**J.N.N**.: Investigation; Writing (review and editing).

**K.M**.: Investigation (intervention delivery); Writing (review and editing).

**A.Z**.: Data analysis and interpretation

**W.D**.: Investigation (intervention delivery); Writing (review and editing).

**W.Z.R**.: Conceptualization; Supervision; Writing (review and editing).

**A.J.N**.: Resources (blinded neuroradiology review of baseline and follow-up MRI); Writing (review and editing). **B.A.C**.: Investigation (neurological assessment, EDSS); Resources (recruitment through neurology clinics); Writing (review and editing).

**M.G.B**.: Conceptualization (neuroimaging); Methodology; Funding acquisition; Supervision; Writing (review and editing).

**M.S.S**.: Conceptualization; Methodology; Funding acquisition; Supervision; Project administration; Writing (review and editing).

All authors read and approved the final manuscript.

## Data availability

The de-identified data that support the findings of this study are available from the corresponding author on reasonable request.

## Ethics approval and consent to participate

The study was approved by the Northwestern University Institutional Review Board (STU00219535) and registered at ClinicalTrials.gov (NCT06276634) before enrollment began. All participants provided written informed consent before participation.

